# Kinetics and specificity of orthoflaviviruses NS1 IgG after Zika virus infection in Yellow fever and dengue immunized individuals: a memory recall hypothesis

**DOI:** 10.64898/2026.09.10.26362723

**Authors:** Solène Marquine, Nazli Ayhan, Kelly Meneyrol, Erwan Le Dault, Sarah Attoumani, Elif Nurtop, Gilda Grard, Aurélie Trignol, Bruno Coutard, Marie Mura, Sébastien Briolant, Cyril Badaut

## Abstract

Pre-existing *Orthoflavivirus* immunity can shape antibody recall upon infection with a heterologous virus. We monitored Zika virus (ZIKV) infection in individuals vaccinated against yellow fever virus (YFV), with or without prior dengue virus (DENV) exposure. We obtained longitudinal serology data (NS1- and whole-virus IgG responses) and neutralization titers on days 0 and 28 after ZIKV infection. YFV-neutralizing antibody levels remained stable over time, consistent with the long-lasting immunity induced by YFV vaccination. By contrast, ZIKV infection triggered an increase in anti-YFV NS1 IgG, suggesting a selective recall of YFV-specific memory B cells. A similar trend was observed for anti-DENV NS1 IgG exclusively in DENV-immune individuals. Thus, prior *Orthoflavivirus* exposure shapes the specificity and magnitude of NS1-directed humoral recall, with implications for serological interpretation.

## Introduction

The yellow fever virus (YFV) belongs to the genus *Orthoflavivirus* within the family *Flaviviridae*. This genus includes several other human pathogens, such as dengue virus (DENV), Japanese encephalitis virus (JEV), West Nile virus (WNV), and Zika virus (ZIKV). YFV is endemic in parts of sub-Saharan Africa, as well as in Central and South America, but remains absent from Asia ^1^. According to the World Health Organization (WHO), YFV causes about 200,000 infections and 30,000 deaths annually ^2^. By contrast, DENV infects an estimated 390 million individuals each year, and cause approximatively 20,000 deaths annually ^3^. ZIKV, first identified in Uganda in 1947, has recently caused large-scale outbreaks in French Polynesia and the Americas ^4^.

Infections caused by these orthoflaviviruses can lead to severe clinical manifestations. DENV and YFV infections may cause hemorrhagic syndromes, whereas, in rare cases, ZIKV infection can result in neurological complications, such as fetal microcephaly and Guillain-Barré syndrome ^5,6^. One recent prevalence study highlighted the extensive geographic overlap and global cocirculation of YFV, DENV, and ZIKV and estimated that 5.6 billion people were at risk of infection with DENV and other arboviruses ^7^. In many regions in which these viruses co-circulate, individuals are often repeatedly exposed to different orthoflaviviruses during the course of their lifetime. These arboviruses have a number of antigenic features in common; natural infection and vaccination therefore generate broadly cross-reactive humoral responses ^8^, raising important questions about the ways in which prior immunity shapes subsequent antibody responses, with important implications for diagnosis and future vaccine strategies.

Non-structural protein 1 (NS1) plays a key role in the pathogenesis of orthoflaviviruses by contributing to viral replication and to the virus’s ability to evade innate and adaptive immune responses ^9,10^. Anti-NS1 antibodies are functionally ambivalent: they can protect against ZIKV ^11,12^ but may also induce molecular mimicry, leading to the targeting of host antigens or platelets during DENV infection ^13^. The B-cell epitopes of the ZIKV NS1 differ considerably from those of YFV and DENV, resulting in very limited IgG cross-reactivity among the orthoflavivirus NS1 proteins ^14,15^. Most ZIKV NS1-specific monoclonal antibodies (mAbs) fail to recognize DENV or YFV NS1, and serum samples from ZIKV-infected individuals display minimal cross-reactivity with heterologous NS1 antigens. Similarly, following primary DENV infection, anti-NS1 mAbs display specificity for the infecting virus ^16–18^. However, structural studies have revealed the presence of conserved NS1 epitopes recognized by a mAb (1G5.3) binding a highly conserved NS1 patch, cross-reacting with several orthoflaviviruses, including ZIKV, DENV, and YFV, and mitigating NS1-mediated pathogenesis ^19^.

YFV vaccination provides a valuable model for studying cross-reactive memory. Indeed, the live-attenuated 17D YFV vaccine rapidly confers long-lasting protection, often persisting for decades, with an efficacy largely dependent on durable humoral responses and memory B-cell activation ^20^. Sequential infections with DENV and ZIKV highlight the complexity of immunity to orthoflaviviruses, whereby heterologous immune responses can modulate disease severity or provide transient protection ^11,21,22^. Oliveira *et al.* suggested that YFV vaccination in ZIKV-infected pregnant women is correlated with protection against congenital microcephaly ^22^. However, other studies have suggested that YFV vaccination is weakly associated with protection against acute ZIKV infection ^23^.

Building on the findings of previous studies, we hypothesize that the memory B cells against NS1 induced by YFV vaccination may be reactivated during subsequent ZIKV infection. Thus, given the pathophysiological role of the NS1 protein, anti-NS1 IgG antibodies may provide transient cross-protection against other orthoflaviviruses for which the patient harbors NS1-directed antibodies. The objectives of this study were to characterize serological dynamics following ZIKV infection in patients with a history of exposure to heterologous orthoflaviviruses. We first assessed the potential contribution of YFV-induced memory B cells to the anti-ZIKV NS1 response in YFV-vaccinated subjects (Vaxinnate cohort). After that, we investigated a cohort of YFV-vaccinated patients who experienced acute ZIKV infection with concomitant increases in the levels of anti-ZIKV NS1 IgG and anti-YFV NS1 IgG (ZIFAG cohort). An additional increase in the levels of DENV NS1-specific IgG was also detected in individuals with a history of prior DENV infection.

## Materials and methods

### Ethical approval

At the French National Reference Center for Arboviruses, all blood samples analyzed in this study were first subjected to routine arbovirus diagnosis procedures. In accordance with national regulations, patients were informed of the potential use of residual material for research and of their right to object. Only samples from individuals who expressed no objection (“non-opposition”) were included. All samples were anonymized and connected to files containing information about the date of symptom onset and a written statement of non-objection to use for technical development or diagnostic comparison. The competent ethics committee (*Comité de Protection des Personnes Sud Méditerranée*) approved the study “*Étude descriptive prospective de la maladie à virus Zika au sein de la communauté de défense des Forces Armées en Guyane (ZIFAG)”,* which was registered on February 29, 2016 as RCB: 2016-A00394-47. Written informed consent was obtained from all patients and the appropriate institutional forms were archived. The Vaxinnate cohort was approved by the competent ethics committee (*Comité de Protection des Personnes Est IV*, reference 2023PPRC01; IDRCB: 2023-A01356-39) on September 25, 2023. Written informed consent was obtained from all participants before inclusion. All research was performed in accordance with relevant guidelines and the Declaration of Helsinki.

### Clinical samples

Clinical samples were obtained from two cohorts. A prospective one-year cohort study (**ZIFAG**) of ZIKV-infected patients was conducted in French Guiana during the 2016–2017 ZIKV epidemic ^24^. ZIKV infection was confirmed by RT-PCR. Multiple serum samples were collected from 34 patients over a one-year period following the onset of Zika symptoms. Samples were harvested at seven to 11 time points: at 3, 5, 7, 14, 21 and 28 days post infection, and at months 2, 3, 6, 9 and 12. All patients were vaccinated against YFV before their arrival in French Guiana. In addition, some patients were tested positive for previous DENV infection by serology. Samples from a second cohort (**Vaxinnate**) were also analyzed. This cohort included individuals orthoflavivirus-naïve vaccinated against YFV on the first day of the study. Blood samples were collected on day 1 (pre-vaccination) and eight months after vaccination. In total, 10 paired samples were available for analysis at these two time points.

Moreover, a panel of arbovirus-negative serum samples was used to define the positivity threshold for the Luminex DENV NS1 IgG and ZIKV NS1 IgG (*n* = 110), and YFV NS1 IgG (*n* = 33) tests. Test sensitivity was evaluated with 83 DENV-positive serum samples collected before 2014 and confirmed by seroneutralization, 46 ZIKV-positive sera collected during the late phase of infection and confirmed either by PCR on an acute-phase serum sample or by a ZIKV-specific seroneutralization assay, 4 serum samples from individuals with YFV infection confirmed by YFV seroneutralization and 10 serum samples from YFV-vaccinated individuals. All samples were provided by the French National Reference Center for Arboviruses.

### Production and purification of NS1 antigens and viruses for ZIKV, YFV, and DENV1-4

NS1 proteins from YFV (UniProtKB/Swiss-Prot: P03314.1, yellow fever virus 17D) and DENV-4 (UniProtKB/Swiss-Prot: P09866.2, Dengue virus 4 Dominica/814669/1981) were produced by Bio-Techne (Minneapolis, USA). Recombinant NS1 proteins from DENV-1, ‡2 and ‡3 and ZIKV were produced and purified in the laboratory according to the following protocol. Human embryonic kidney (HEK) cells were transfected with plasmid DNA carrying the NS1 protein-encoding gene. The pcDNA3.1 expression plasmid was assembled to contain, in order: a Kozak consensus sequence, the tissue plasminogen activator (tPA) signal peptide to ensure secretion, the NS1 coding sequence, a 6×His C-terminal tag, and a stop codon. The construct was inserted into the pcDNA3.1 backbone carrying the ampicillin resistance (AmpR) marker. HEK Expi293™ cells (Thermo Fisher Scientific) were cultured at a concentration of 4×10^6^ cells/mL in Expi293™ expression medium (ref A1435101), maintained at 37°C under an atmosphere containing 8% CO_2_, at 80% humidity, with shaking at 125 rpm. One hour before transfection the cells were diluted to a density of 1×10^6^ cells/mL and transfected with the kit supplied with the cells (Thermo Fisher Scientific, Expi293™ Expression System Kit, ref. A14635). Four days after transfection, the transfected cell culture was centrifuged for 15 minutes at 4,000 x *g*. The supernatant was removed, mixed with 500 µL Ni-IMAC beads (Chelating Sepharose fast-flow, Cytiva, ref. 17057502) previously equilibrated with a buffer consisting of 50 mM Tris HCl pH 8 and 300 mM NaCl, and incubated for 2 hours at 4°C with shaking. Elution was performed with the equilibration buffer supplemented with 100 mM imidazole. Two successive dialyses were performed against 1 X PBS supplemented with 350 mM NaCl, at 4°C. Protein concentration was evaluated by spectrophotometry at 280 nm and purity was evaluated by SDS-PAGE with Coomassie blue staining. The protein preparation was split into aliquots and stored at ‡80°C.

The following viruses were produced: ZIKV (French Polynesia, 2013, GenBank accession number KJ776791), DENV1 (DENV1_CNR-SN_VCT_2012, GenBank accession number PP695350), DENV2 (Martinique DENV2 98-703 strain 1998, GenBank accession number AF208496), DENV3 (Martinique DENV3, GenBank accession number AH011666), DENV4 (DENV4_CNR-SN-IRBA-814_IDN_2014, GenBank accession number PP695349) and YFV (GenBank accession number MF405338). Vero cells were exposed to 0.01 MOI of ZIKV, DENV1-4 or YFV and cultured in Dulbecco’s modified Eagle’s medium (DMEM) supplemented with 2% heat-inactivated fetal bovine serum (FBS) at 37°C, under an atmosphere containing 5% CO_2_, for 3.5 days (ZIKV), 7 days (DENV) or 4-5 days (YFV). After incubation, the culture supernatants were collected and the viral particles precipitated with polyethylene glycol 6000 (10% w/v PEG 6000) and 1 M NaCl. The precipitates obtained were washed and resuspended in PBS supplemented with 7.5 mM HEPES (pH 8).

### IgG response of ZIKV-infected patients to the NS1 protein of the orthoflaviviruses studied

All inactivated serum samples were analyzed with Luminex multiplex immunoassay (xMAP) technology. The NS1 proteins from ZIKV, YFV and DENV1-4 were covalently coupled to carboxyl-functionalized fluorescent polystyrene beads (BPLX MAG COOH, Luminex. Inc) with the Bio-Plex amine coupling kit (Bio-Rad Laboratories), in accordance with the manufacturer’s instructions. Unreacted sites were blocked with blocking buffer from the amine coupling kit (Bio-Rad Laboratories). Protein-coupled microsphere preparations were subjected to counting in a hemocytometer and were stored in the dark at 4°C. Before use, the coupled beads were vortexed (for 30 s), sonicated (for 30 s) and diluted to 2,000 beads/µL in dilution buffer (Kit EI 2668-9601 G, Euroimmun).

The quality of microsphere coupling was validated with a commercial mouse monoclonal antibody targeting the NS1 protein of DENV-2 and cross-reacting with other DENV serotypes and ZIKV (ab214337, ABCAM). This control assay was performed in a white, low-adsorption, round-bottom polypropylene 96-well plate (Nunc Microwell 267350 Thermo Fisher Scientific). The microsphere mixture was prepared at a final concentration of 20 microspheres per µL, and 50 μL of this mixture was dispensed into each well. The beads mixture was incubated with 50 μL serial dilutions of monoclonal antibody (ab214337, ABCAM) at concentrations of 1000 ng to 0.06 ng/well in wash buffer, for 60 min at room temperature in the dark on a plate shaker (800 rpm). The wash buffer consisted of phosphate-buffered saline (PBS), 3% skim milk and 0.1% Tween-20 (Sigma-Aldrich). The plate was washed with 100 µL wash buffer and placed on a magnetic stand. The solution was discarded. The beads were washed twice in wash buffer and incubated with 50 μL/well secondary antibody (R-Phycoerythrin AffiniPure F(ab’)^D^ Fragment Donkey Anti-Mouse IgG (H+L), part no. 715-116-151, Jackson Immuno Research Europe Ltd) at a concentration of 2.5 µg/mL for 1 hour at room temperature in the dark, with shaking (800 rpm). The plates were washed twice with wash buffer and the beads were resuspended in 100 µL dilution buffer/well and analyzed with a MAGPIX system (Luminex Corp, TX USA). For each set of beads, at least 100 events were read; results are expressed as the median fluorescence intensity (MFI) per 100 beads.

The serum analysis was adapted from the protocol used to test bead coupling: the beads were incubated with 50 μL of a 1/100 dilution of serum in wash buffer. The final serum dilution was 1/200. We used 50 μL/well of secondary antibody at a concentration of 2.5 µg/mL to detect the human IgG (AffiniPure F(ab’)^₂^ Fragment Goat Anti-Human IgG, Jackson Immuno Research Europe Ltd) present on the target bound to the bead. Finally, a sample was considered positive if the signal obtained exceeded the fixed threshold value (Table 3).

### Microneutralization of viruses by human serum samples

The neutralizing antibodies in each serum sample were titrated by microneutralization in a biosafety level 3 laboratory. After filtration through a filter with 0.22 µm pores, the serum samples were serially diluted in PBS in 96-well plates: from 1:20 to 1:320 for ZIKV and DENV, and up to 1:40960 for YFV. Each dilution (100 µL) was incubated with 100 µL viral supernatant, corresponding to titers of 0.5 TCID_50_ per μL for ZIKV (strain H/PF 2013), DENV-1 (strain DENV1_CNR-SN_VCT_2012), DENV-2 (strain DEN2/H/IMTSSA-MART/98-703), DENV-3 (strain D3/H/IMTSSA-MART/2001/2023), DENV-4 (strain DENV-4/1998/ID814) and YFV (Asibi stain), for one hour at 37 °C. The mixtures were then added to Vero cells (ATCC CCL-81, 5 × 10L cells/well). After four days for ZIKV, seven days for DENV, and four to five days for YFV, images of all the wells were obtained with the Incucyte SX5 live-cell analysis system (Sartorius, Göttingen, Germany) and cytopathic effects were evaluated. The neutralizing titer was defined as the highest serum dilution inhibiting any cytopathic effect (CPE). A neutralizing antibody titer ≥ 1:40 was considered positive.

### Statistical and mathematical analysis

Neutralizing antibody titers and Luminex-based IgG ELISA values in the ZIFAG cohort did not follow a normal distribution, as assessed by the Shapiro-Wilk test. Given the non-normality of the data, non-parametric statistical analyses were performed to evaluate differences in the NS1-specific IgG responses measured with Luminex technology. Kruskal-Wallis tests were performed to assess overall differences between groups, followed by Dunn’s post-hoc tests for multiple comparisons between individuals with and without prior DENV infection. Curves modeling anti-NS1 IgG responses from aggregated patient data were obtained with Wood’s equation ^25,26^ (Kaleidagraph 4.5). Mann-Whitney tests were used to assess differences between groups with and without memory responses. Serum neutralization on days 0 and 28 was compared in a Wilcoxon test (GraphPad Prism, v10.2).

### Sequence alignment, and structural study

The NS1 protein sequences of YFV, DENV-1 to ‡4 and ZIKV were aligned in Geneious R11 software, with the plugin ‘muscle’ and the BLOSUM62 matrix, using a threshold of 0. Protein structures were aligned, visualized and analyzed with PyMol (V 3.1.6.1).

## Results

### Reliability of the Luminex test for IgG against ZIKV-, DENV- or YFV-NS1

Positivity thresholds for the Luminex assay were determined with negative control serum samples for *Orthoflavivirus* infection (negative PCR and IgM/IgG serology). Thresholds were calculated as the mean fluorescence intensity (MFI) plus three standard deviations for 110 serum samples for anti-ZIKV and anti-DENV1–4 NS1 IgG, and 33 serum samples for anti-YFV NS1 IgG. The resulting cutoffs were 679 MFI (ZIKV), 427 MFI (DENV1), 1802 MFI (DENV2), 207 MFI (DENV3), 453 MFI (DENV4), and 903 MFI (YFV) (Supplementary Table 1).

Luminex assay sensitivity was assessed with serum samples from patients infected with ZIKV (*n* = 46; 100% sensitivity) or DENV (*n* = 83; 98% sensitivity). For YFV, sensitivity (*n* = 4 + 10; 100%) was determined with samples from four YFV-infected patients and 10 individuals vaccinated against YFV from the Vaxinnate cohort (Table 1) The vaccinated individuals had no history of travel to regions endemic for DENV, ZIKV, or YFV.

**Table 1.**
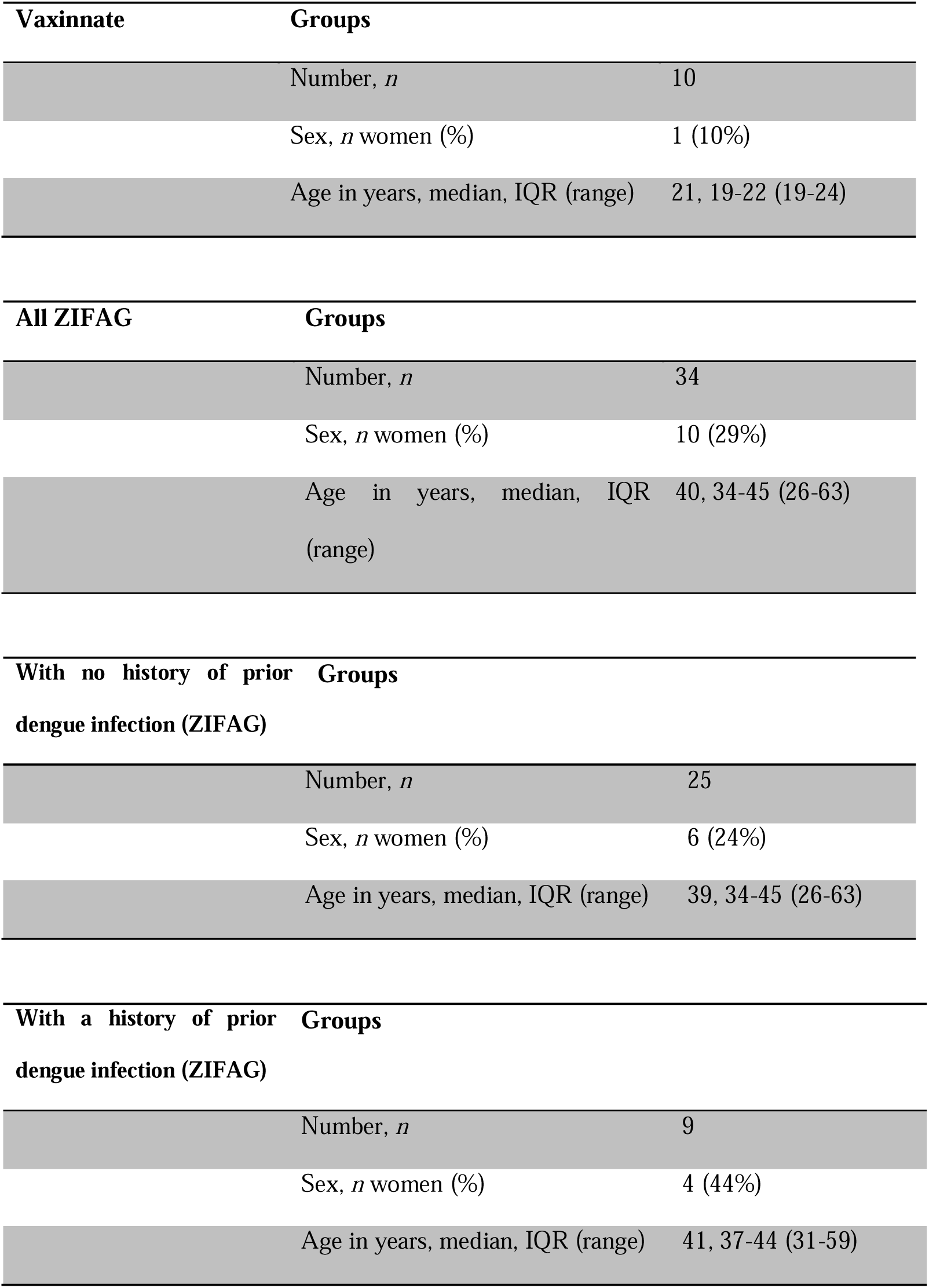
Description of the groups from the ZIFAG and Vaxinnate cohorts.

**Table 2.** Fluorescence intensity of anti-NS1 IgG for DENV1 to DENV4, ZIKV and YFV on day 28, in serum samples from patients with and without a history of prior DENV infection in ZIFAG cohort. This table is linked to figure 2.

|  | DENV1 | DENV2 | DENV3 | DENV4 | ZIKV | YFV | DENV1 NS1 | DENV2 NS1 | DENV3 NS1 | DENV4 NS1 | ZIKV NS1 | YFV NS1 |
| --- | --- | --- | --- | --- | --- | --- | --- | --- | --- | --- | --- | --- |
|  | NS1 | NS1 | NS1 | NS1 | NS1 | NS1 | pr. DENV | pr. DENV | pr. DENV | pr. DENV | pr. DENV | pr. DENV |
| **Number of values** | 25 | 25 | 25 | 25 | 25 | 25 | 9 | 9 | 9 | 9 | 9 | 9 |
| **Minimum** | 131 | 213 | 44 | 72 | 5487 | 523 | 205 | 323 | 79 | 184 | 6877 | 1049 |
| **25% Percentile** | 216 | 398.5 | 73.5 | 159.5 | 7471 | 1490 | 212.5 | 481 | 110.5 | 286.5 | 8051 | 1315 |
| **Median** | 371 | 533 | 136 | 266 | 8684 | 4067 | 572 | 928 | 165 | 488 | 9532 | 1962 |
| **75% Percentile** | 960 | 1407 | 356 | 860.5 | 10399 | 6711 | 3092 | 4655 | 2579 | 7104 | 12380 | 4939 |
| **Maximum** | 4120 | 5661 | 4172 | 3286 | 15007 | 16181 | 14883 | 18024 | 15042 | 12196 | 14786 | 11876 |
| **Range** | 3989 | 5448 | 4128 | 3214 | 9520 | 15658 | 14678 | 17701 | 14963 | 12012 | 7909 | 10827 |
| **10% Percentile** | 158.4 | 310.2 | 51.2 | 78.8 | 6284 | 692.8 | 205 | 323 | 79 | 184 | 6877 | 1049 |
| **90% Percentile** | 2140 | 3023 | 1654 | 1339 | 13232 | 10284 | 14883 | 18024 | 15042 | 12196 | 14786 | 11876 |
| **Threshold** | 427 | 1802 | 207 | 453 | 679 | 903 | 427 | 1802 | 207 | 453 | 679 | 903 |
| **Positives** | 12 | 3 | 9 | 7 | 25 | 22 | 5 | 3 | 3 | 5 | 9 | 9 |
| **% positives** | 48 | 12 | 36 | 28 | 100 | 88 | 56 | 33 | 33 | 56 | 100 | 100 |

The high sensitivity of the Luminex assay against the three tested flaviviruses NS1 protein allows us to question the specificity and cross-reactivity of the human immune response.

### ZIKV infection triggers increased IgG responses to heterologous *Orthoflavivirus* NS1 proteins in prior exposure-dependent manner

In total, 34 individuals infected with ZIKV were included in the ZIFAG cohort study: 29% were women and 71% were men (Table 1). Median age in these individuals was 40 years, with age ranged from 26 to 63 years. The selected patients were assigned to two groups. The first group included patients who had never had an *Orthoflavivirus* infection i.e., the first serum sample (day < 4) contained no total detectable IgG against orthoflaviviruses (*n* = 25) (Table 1) ^27^. This group comprised 24% women and 76% men, with a median age of 39 years, (range: 26-63 years). The second group included patients with a history of prior DENV infection (*n* = 9), i.e., the first sample already contained total IgG against orthoflaviviruses in addition to neutralizing antibodies against DENV (Table 1). This second group comprised 44% women and 56% men, aged from 31 to 59 years, with a median age of 41 years.

IgG responses to the NS1 proteins of ZIKV, YFV, and DENV1-4 were then evaluated in individuals with (*n* = 9) or without (*n* = 25) a history of prior DENV infection. For the analysis of antibody kinetics, we first applied a mathematical model to the aggregated longitudinal data (Supplementary Table 2). In parallel, we used IgG levels measured 28 days after symptom onset as an indicator of maximum response (Figure 1), taking into account the variability of individual profiles. In individuals who had never been infected with DENV, anti-ZIKV NS1 and anti-YFV NS1 IgG responses increased rapidly and exceeded the positivity threshold. The modeled kinetics revealed peak responses of 12,016 MFI for anti-ZIKV NS1 IgG on day 118, and 4,531 MFI for anti-YFV NS1 IgG on day 41. By contrast, anti-DENV1-4 NS1 IgG levels remained below the detection threshold throughout the study. In participants with a history of prior DENV infection, IgG responses to ZIKV-NS1 and YFV-NS1 followed similar kinetics, reaching 12,911 and 3,432 MFI on days 101 and 49, respectively. However, unlike the DENV-naïve group, these individuals also developed strong IgG responses to DENV1-4 NS1 proteins, with peak MFIs of 3,195 (DENV1), 4,556 (DENV2), 2,484 (DENV3), and 3,366 (DENV4) occurring between days 72 and 89 post-ZIKV infection.

**Figure 1.**
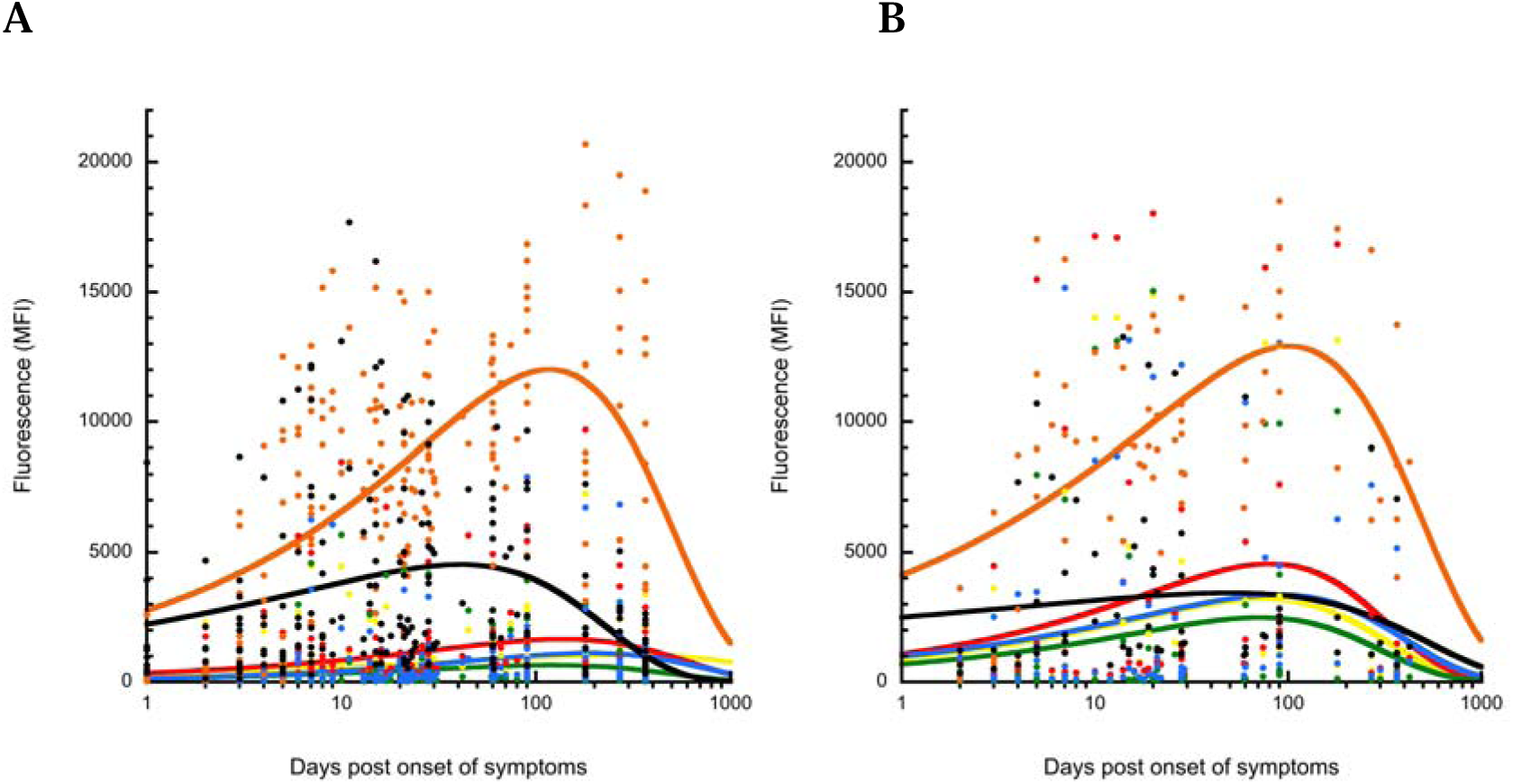
Modeling of anti-NS1 IgG kinetics for ZIFAG cohort. **A**: Patients with no history of prior DENV infection **B**: Patients with a history of prior DENV infection. Corresponding curves: orange: ZIKV NS1; gray: YFV NS1; yellow: DENV1 NS1; red: DENV2 NS1; green: DENV3 NS1; blue: DENV4 NS1. The threshold values for this test - which detects IgG through a multiplex technique targeting the NS1 proteins of YFV, ZIKV and DENV (1 to 4) - were 903, 679, 427, 1802, 207, and 453 MFI, respectively.

ZIKV infection was associated with increased modeled IgG responses to ZIKV and YFV NS1 in all participants, whereas IgG responses to DENV1-4 NS1 were observed only in individuals with prior DENV infection.

In addition to the modeling approach, we compared anti-NS1 IgG levels on day 28 after symptom onset between individuals with and without prior DENV infection in the ZIFAG cohort (Supplementary Figure 1). As expected, IgG levels on day 2 were low and below the detection thresholds in both groups.

Within the DENV-naïve group, median MFIs for IgG against the NS1 proteins of DENV1, DENV2, DENV3, DENV4, ZIKV, and YFV were 371, 533, 136, 266, 8684, and 4067, respectively. In these individuals, IgG responses to the NS1 proteins of ZIKV and YFV were significantly stronger than those to all four DENV serotypes (p < 0.001 and p = 0.002, respectively; Figure 2).

**Figure 2.**
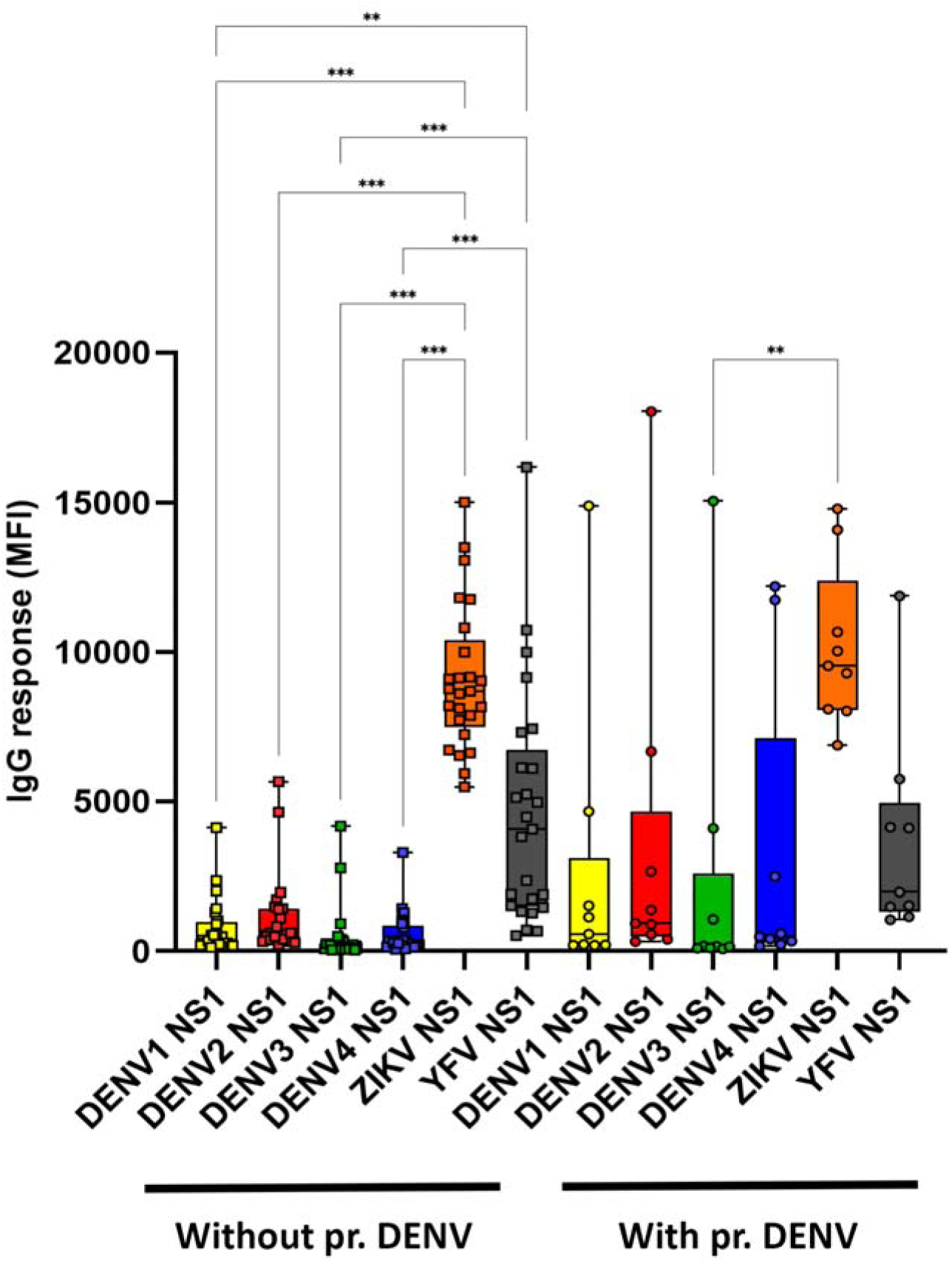
Intensity of the IgG response to the NS1 proteins of DENV1-4, ZIKV, and YFV, 28 days after ZIKV infection in ZIFAG cohort. Mean fluorescence intensities (MFIs) were compared between serum samples from patients with (“pr. DENV”) and without a history of prior dengue infection. Boxes represent the 10th to 90th percentiles, with whiskers extending from the minimum to the maximum, and the median indicated at the center. The threshold values for this test - which detects IgG through a multiplex technique targeting the NS1 proteins of YFV, ZIKV and DENV (1 to 4) - were 903, 679, 427, 1802, 207, and 453 MFI, respectively.

In the prior-DENV group, median MFIs were 572, 928, 165, 488, 9532, and 1962 for DENV1, DENV2, DENV3, DENV4, ZIKV, and YFV, respectively. Within this prior-DENV group, the IgG response to ZIKV NS1 was significantly higher than the response to DENV3 NS1 (p = 0.0013), whereas no other significant differences were observed.

When comparing the two groups of the ZIFAG cohort, although individuals with a history of prior DENV infection displayed a trend toward higher median MFI values for DENV1-4 NS1, these differences did not reach statistical significance. Similarly, IgG responses to ZIKV and YFV NS1 were comparable between the two groups (p > 0.99 for both).

### No cross-reactive anti-NS1 IgG detection in YFV-vaccinated individuals

We previously characterized the immune response induced by ZIKV infection in YFV-vaccinated individuals in more detail by next determining the cross-reactivity of anti-YFV NS1 IgG with ZIKV-NS1. Here, in the Vaxinnate cohort of *Orthoflavivirus* naïve individuals vaccinated against YFV with the live attenuated 17D strain, we observed a significant increase in anti-YFV-NS1 IgG levels between the day of vaccination (D1) and 8 months after vaccination (M8) (*n* = 10, Šídák’s multiple comparisons test, *p* = 0.02). By contrast, no significant change in anti-ZIKV NS1 IgG levels was observed over the same period (Figure 3). YFV vaccination induced a specific anti-YFV NS1 IgG response without detectable cross-reactive IgG responses to ZIKV NS1.

**Figure 3.**
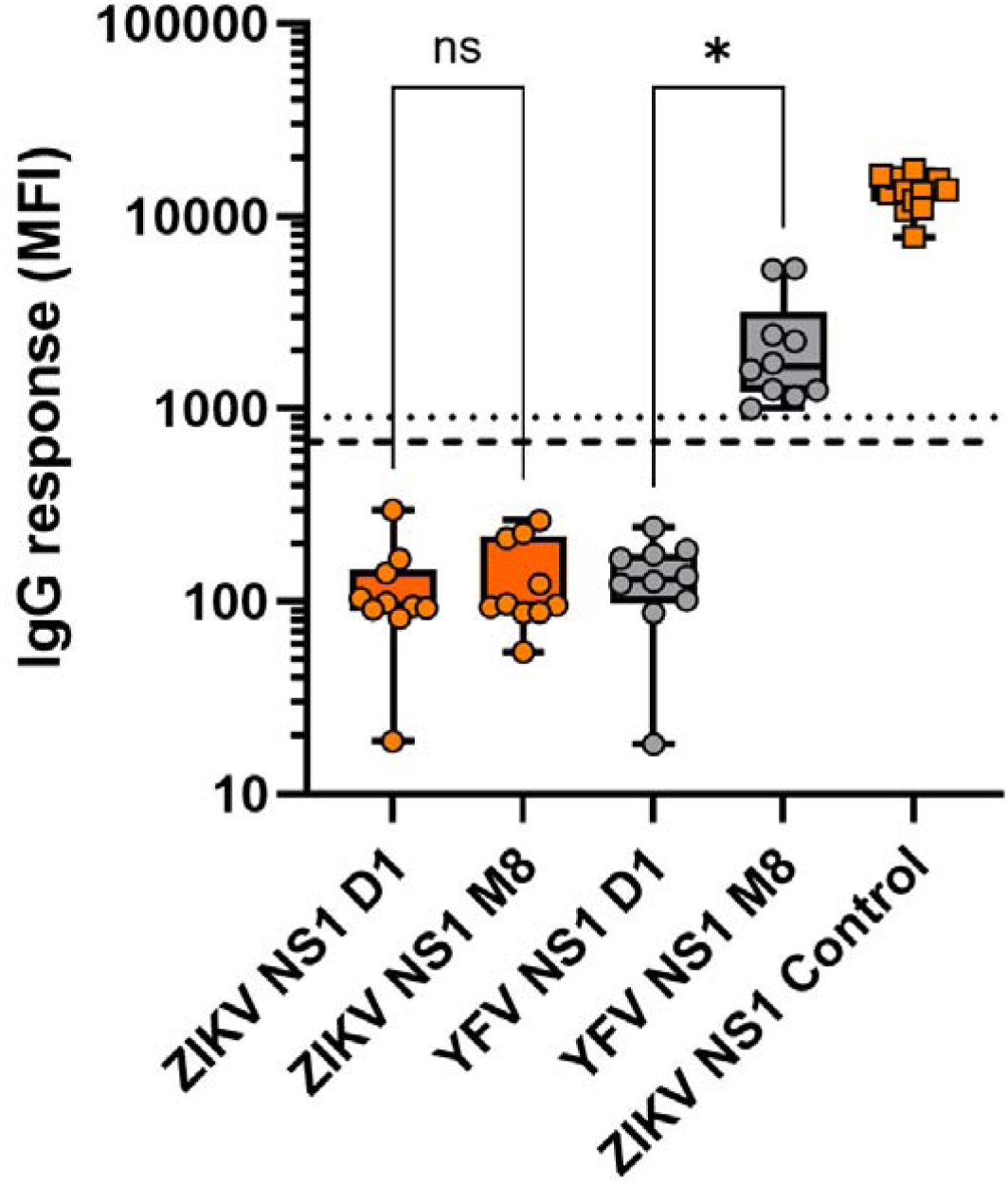
Absence of cross-reactivity between IgG directed against the NS1 proteins of ZIKV and YFV in Vaxinnate cohort. Anti-ZIKV NS1 IgG (orange) and anti-YFV NS1 IgG (gray). IgG responses are shown on day 1 (D1) and at 8 months (M8) after YFV vaccination (*n* = 10). The “Control” consists of ZIKV-positive patients (*n* = 14). Fluorescence intensity (MFI) values are plotted on the *y*-axis. Whiskers indicate the minimum and maximum values and the center line corresponds to the median. The dotted line represents the positivity threshold for anti-YFV NS1 IgG and the dashed line represents the positivity threshold for anti-ZIKV NS1 IgG. One-way ANOVA (mixed-effects analysis with Geisser-Greenhouse correction) was used for statistical analysis.

### ZIKV infection boosts the production of neutralizing antibodies against DENV but not YFV in DENV-naive individuals from ZIFAG cohort

Antibody neutralization titers on days 0 and 28 post-ZIKV infection were analyzed. The titers of neutralizing antibodies against ZIKV and DENV increased significantly between days 0 and 28 in patients who had never been infected with DENV (median log_2_ ZIKV titer: D0 = 4.32, D28 = 6.32, *n* = 22; W = 180; *p* < 0.001; median log_2_ DENV1 titer: D0 = 4.32, D28 = 7.32, *n* = 22; W = 253; *p* < 0.001, respectively) (Figure 4). By contrast, no change was observed for neutralizing antibody titers against YFV over the same period (median log_2_ titer: D0 = 7.32, D28 = 7.32, n = 23; W = 50; *p* = 0.1536) (Figure 4B).

**Figure 4.**
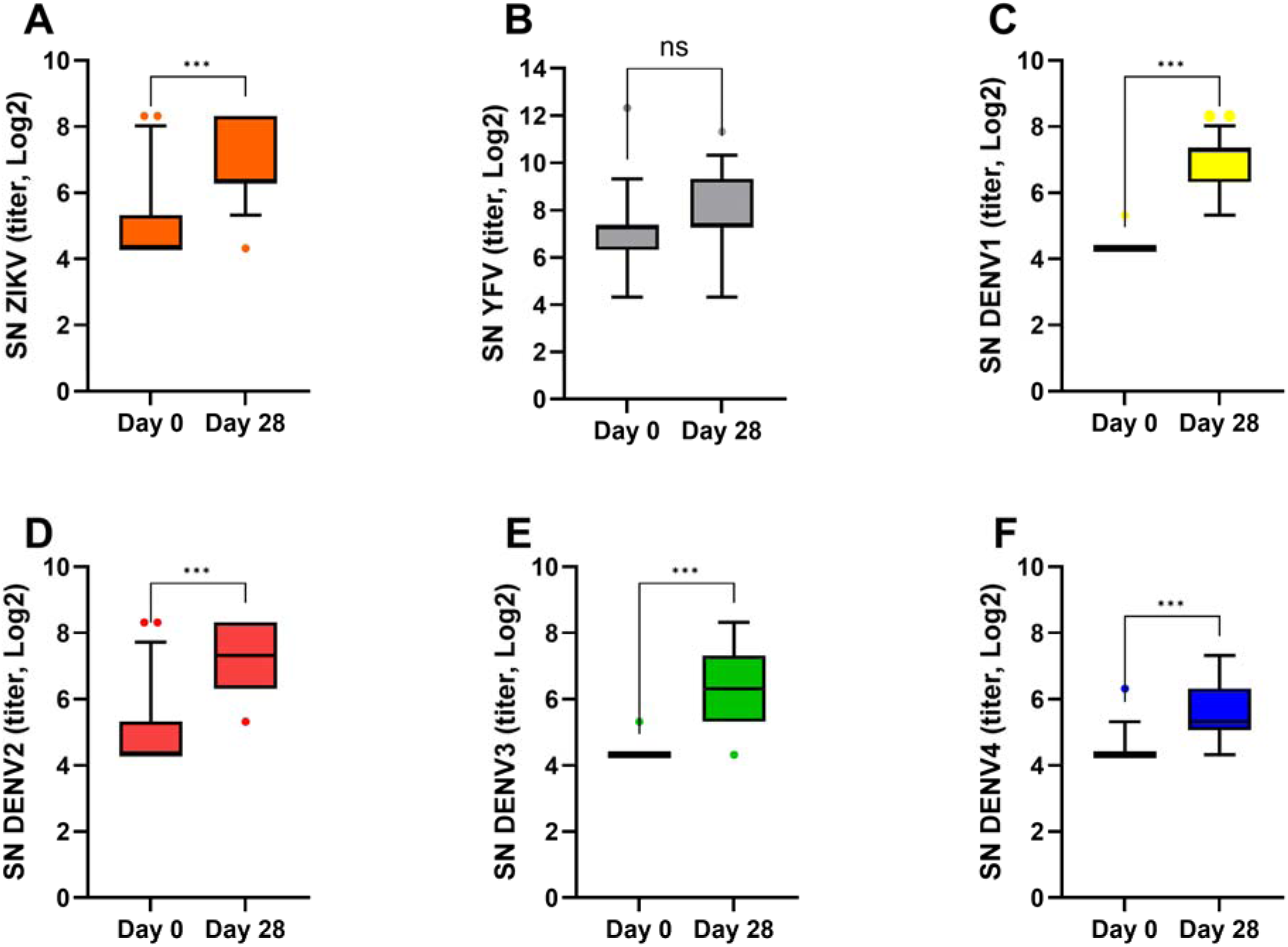
Seroneutralization (SN) measured on Days 0 and 28 for Zika virus (post-Zika infection), yellow fever virus, and the four dengue virus serotypes in patients with no prior history of DENV infection from ZIFAG cohort. Panels: A: ZIKV, B: YFV, C: DENV1, D: DENV2, E: DENV3, F: DENV4. The central line indicates the median, and the whiskers represent the 10th and 90th percentiles.

In patients with a history of prior DENV infection, a significant increase in anti-DENV1 neutralizing antibody titers was observed (median log_2_ titer: D0 = 4.32, D28 = 7.32, *n* = 7; W = 28; *p* = 0.016). Similar results were obtained for DENV3. No significant changes were observed for ZIKV (median log_2_ titer: D0 = 4.32, D28 = 6.32, *n* = 7; W = 12; *p* = 0.281), YFV (median titer: D0 = 6.32 D28 = 7.32, *n* = 8; W = 12; *p* = 0.188) or DENV2 or DENV4 neutralizing antibody titers (Supplementary Figure 2).

ZIKV infection was associated with increases in neutralizing antibody titers against ZIKV and DENV in DENV-naïve individuals, whereas YFV neutralizing antibody titers remained high and stable.

### The low conservation of epitopes carried by the *Orthoflavivirus* NS1 protein suggests limited antibody cross-reactivity

We aimed to assess whether a cross-reactive memory recall mechanism could account for our observations. Accordingly, it was critical to evaluate the presence of shared structural determinants, including both immunodominant and subdominant epitopes. Alignment of the NS1 sequences from the six studied viruses (Figure 5) revealed that the YFV NS1 protein sequence shared 72.7% to 76.1% similarity and 42.3% to 47.0% identity with those of ZIKV and the four DENV serotypes (Table 3).

**Figure 5.**
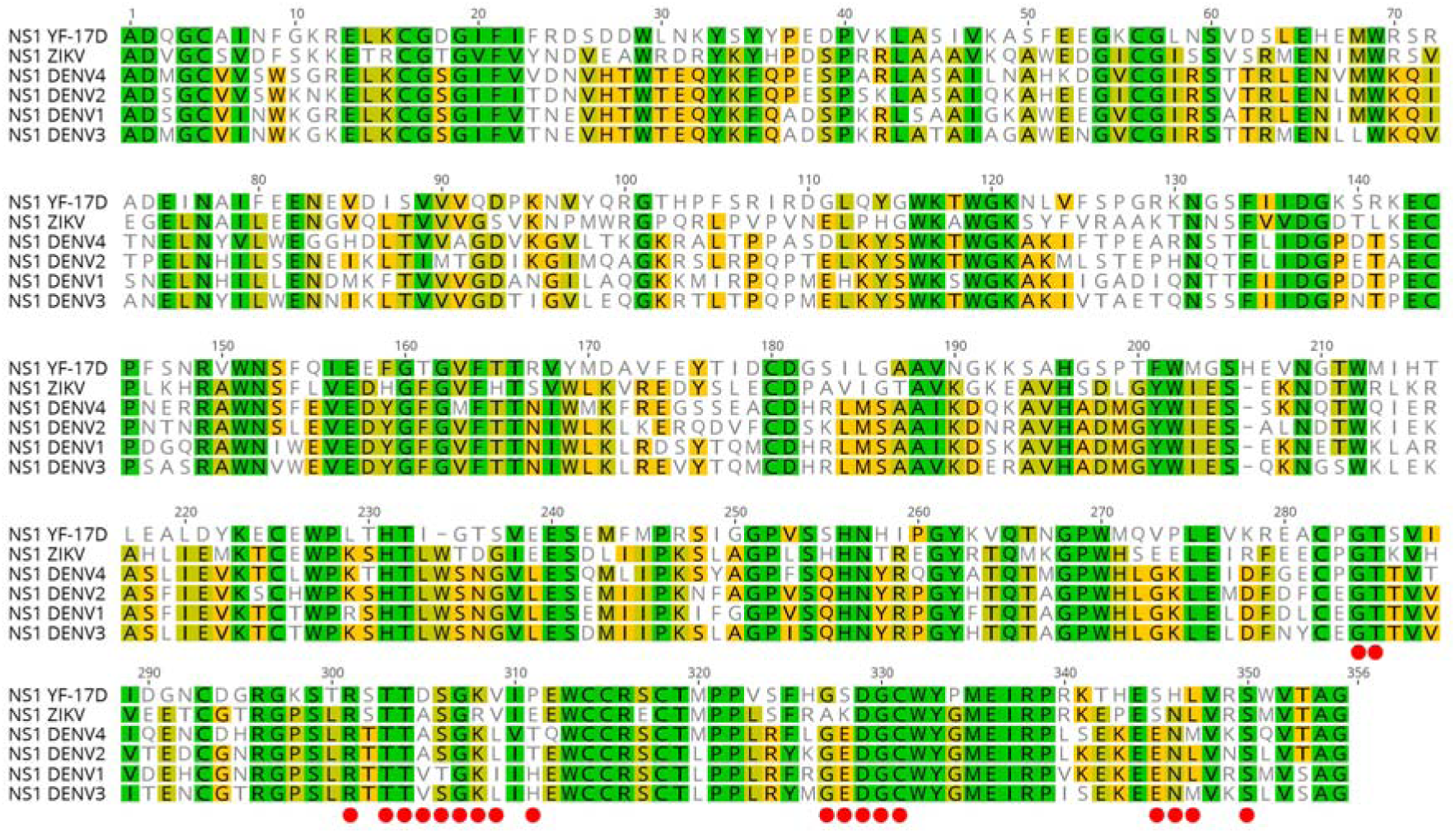
Multiple alignment of NS1 sequences from YFV, ZIKV, and the four DENV serotypes. The NS1 protein sequences of YFV 17D (P03314), ZIKV, Thailand, 2021 (OQ734448); DENV1, France, 2022 (PX442212); DENV2, La Réunion island, 2018, (PX309104), DENV3, Cuba, 2022 (PX442213); DENV4, Dominica, 1981 (UOS86121) were aligned to identify conserved and similar residues. Identical amino acids are shown in dark green, whereas similar amino acids (80-100%, 60-80%, less than 60% similarity according to the Blosum62 matrix, threshold = 0) are shown in light green, yellow, and white, respectively. Residues corresponding to the epitope recognized by the 1G5.3 mAb, identified in the crystallographic structures of the carboxy-terminal domains of the ZIKV NS1 protein (PDB: 7BSD) and DENV2 NS1 (PDB: 7BSC), are indicated by red dots.

**Table 3.** Identity matrix (bottom left) and similarity matrix (top right) between the NS1 proteins from yellow fever virus (YFV), Zika virus (ZIKV) and dengue serotypes (DENV1-4). Color scale: red (minimum) green (maximum).

|  | NS1_YF_<br>17D | NS1_Zika<br>_62136 | NS1_DENV4<br>-UOS86121 | NS1_DENV2<br>_47099 | NS1_DENV1<br>_62902 | NS1_DENV3<br>_62678 |
| --- | --- | --- | --- | --- | --- | --- |
| NS1_YF_17D |  | 76.1 | 73.8 | 74.4 | 72.7 | 73.5 |
| NS1_Zika_62136 | 47.0 |  | 82.5 | 82.5 | 82.2 | 82.2 |
| NS1_DENV4-UOS86121 | 43.7 | 54.2 |  | 91.0 | 89.8 | 90.7 |
| NS1_DENV2_47099 | 43.9 | 53.4 | 72.3 |  | 91.2 | 92.4 |
| NS1_DENV1_62902 | 43.1 | 54.2 | 68.9 | 74.3 |  | 94.9 |
| NS1_DENV3_62678 | 42.3 | 55.9 | 73.7 | 74.6 | 79.7 |  |

A crystallographic structure of complexes formed between a Fab fragment of the 1G5.3 monoclonal antibody and the C-terminal domain of ZIKV NS1 (PDB 7BSD) or DENV2 NS1 (PDB 7BSC) identified a cross-reactive epitope in several orthoflaviviruses ^19^. We performed a structural alignment (Figure 6) of YFV NS1 (blue, accession number 8ZBA) with the carboxy-terminal part of ZIKV NS1 (green) complexed with Fab 1G5.3 (gray), which revealed a perfect spatial alignment of these NS1 structures (RMSD _alpha carbon_ = 1.52 Å) ^28^. The amino acids constituting the carboxy-terminus domain of the ZIKV NS1 epitope (orange) overlapped spatially with and were highly similar to those of the NS1 epitope YFV (dark blue). These similar or identical residues are indicated by red dots in Figure 1 (amino acids 301-311 and 327-311).

**Figure 6.**
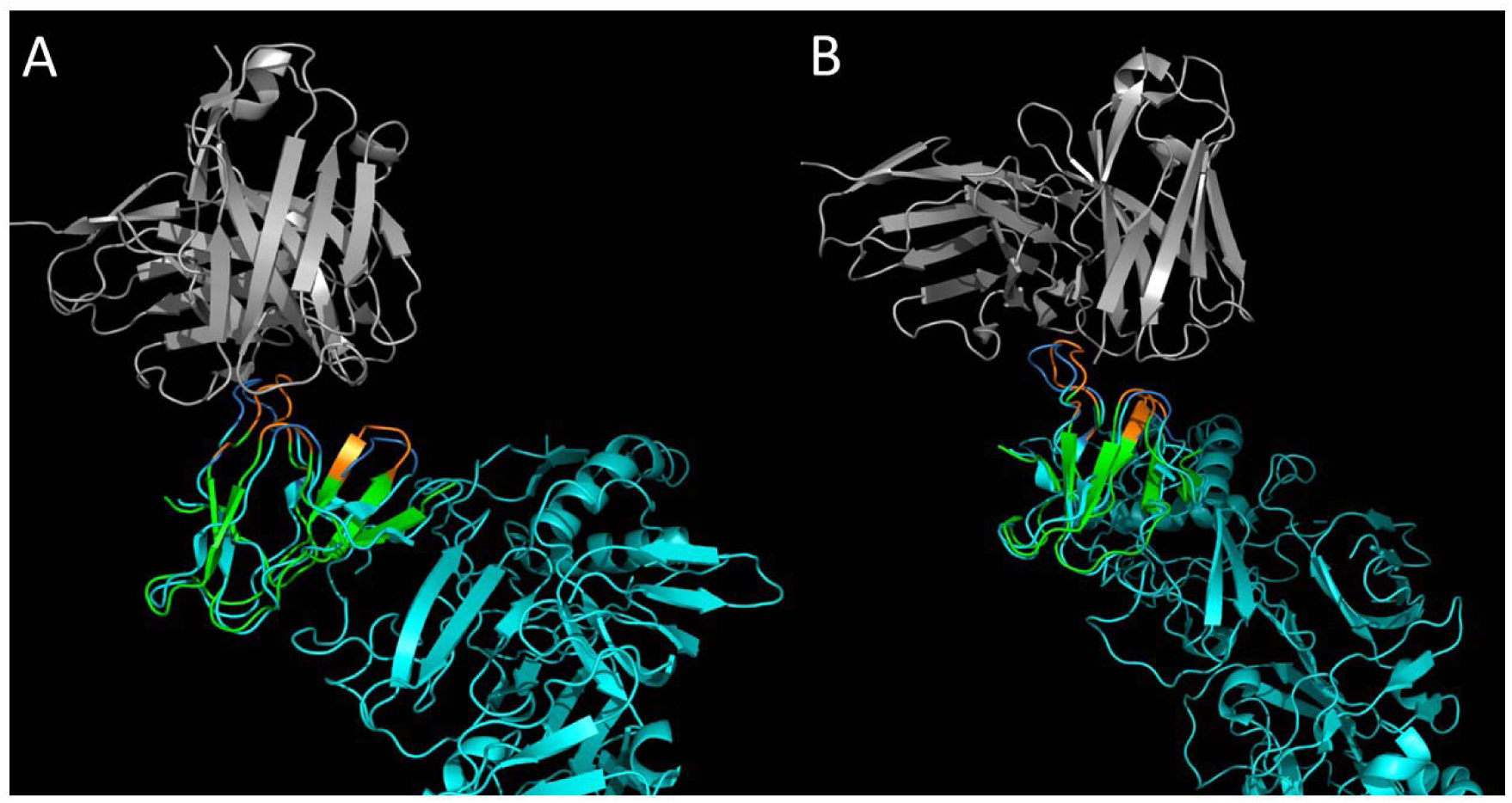
Structural alignment of YFV-NS1 and ZIKV-NS1 in complex with a Fab from a monoclonal antibody displaying cross-reactivity. Two views of the NS1 protein (PDB: 7BSD) from ZIKV (in green) showing an epitope (in orange) recognized by the 1G5.3 antibody (variable region in gray). This epitope is common to the NS1 protein (PDB: 8ZBA) of YFV (in blue): it is spatially aligned and its amino acids are identical or very similar (in dark blue).

Analyses of both sequence and three-dimensional structural homologies of *Orthoflavivirus* NS1 proteins indicated a minimal potential for antibodies cross-reactivity, primarily between ZIKV and YFV NS1.

## Discussion

The level of protection in individuals previously exposed to orthoflaviviruses can be monitored by assessing seroneutralizing antibody responses. Here, we investigated the impact of ZIKV infection in individuals with prior YFV vaccination and, in a prior DENV infection. We monitored changes in neutralizing antibody responses to ZIKV during the early phase of infection. YFV vaccination is known to confer long-lasting protection through the induction of neutralizing antibodies, the presence of which is strongly correlated with protective immunity ^29,30^. Neutralizing antibodies against YFV have been shown to be stable over the long term, with a minimal decline over time ^2,31,32^. In this study, exposure to a heterologous *Orthoflavivirus* did not significantly alter the YFV-neutralizing antibody titers measured between days 0 and 28 post-ZIKV infection. This stability probably reflects the presence of high levels of pre-existing YFV-induced antibodies and is consistent with previous studies demonstrating that a single dose of YFV vaccine can induce long-term, often lifelong, immune responses within 14 days post-vaccination ^33^. In individuals with a history of prior DENV infection, a DENV-neutralizing response was already present at the time of ZIKV infection (day 0), as previously observed ^34^.

In addition to serum-neutralizing antibodies that provide direct antiviral protection, antibodies targeting the NS1 protein of orthoflaviviruses have attracted considerable interest due to the protein’s involvement in the pathogenesis of infection. Anti-NS1 antibodies have been shown to confer or correlate with protection against several orthoflaviviruses, including dengue, West Nile, Zika, and yellow fever viruses, as recently highlighted by Tamietti et al., together with previous experimental studies (^12,19,35,36^). However, their degree of cross-reactivity remains an important consideration, as it may influence their capacity to mediate cross-protective immunity against diseases caused by related orthoflaviviruses. An increase in IgG antibodies directed against the NS1 protein of a virus that has never infected the individual would be indicative of antibody cross-reactivity rather than a recall response. This distinction underlies that memory-recall hypothesis is proposed exclusively for previously exposed individuals (YFV-vaccinated, or prior DENV-infected), and not for naïve participants. By analyzing individuals from the Vaxinnate cohort - who have no history of prior *Orthoflavivirus* exposure - and from the DENV-naïve individuals of the ZIFAG cohort, we confirm that NS1 IgG response was mainly virus-specific, with limited detected cross-reactivity using Luminex technology against NS1 from another *Orthoflavivirus* in a naïve individual. Nevertheless, in line with the presence of shared epitopes between the two viral NS1 proteins, individuals with previous YFV-17D exposure in the ZIFAG cohort exhibited a marked, concurrent increase in both anti-YFV and anti-ZIKV NS1 IgG levels ^37^. A similar pattern was observed for anti-DENV NS1 IgG, exclusively in individuals with prior DENV exposure. This selective rise of NS1 IgG, relying on previous viral exposure only, are strongly in favor of a recall of cross-reactive memory B cells based on subdominant epitopes, rather than genuine cross-reactivity of major epitopes induced by ZIKV NS1 protein exposure during infection. NS1-specific antibodies do not contribute to virus neutralization because NS1 is not present on mature virions, Therefore, NS1 IgG are considered non-cross-neutralizing and, largely non-cross-reactive ^35,38^ but they may protect against NS1-mediated pathogenicity, as reported for several orthoflaviviruses ^39^.

A comparative analysis of NS1 sequences showed only moderate similarity between YFV, ZIKV, and DENV, with few conserved residues, consistent with the limited IgG cross-reactivity observed experimentally and with previous studies. Seven linear B-cell epitopes were identified in poorly conserved regions of the NS1 protein sequences ^40^. Nevertheless, crystallographic data enabled the identification of a common epitope that had not yet been classified as a major epitope ^41^. The subnanomolar affinities of mAb 1G5.3 for ZIKV (*K_d_* = 0.24 nM), YFV (*K_d_* = 2.69 nM), and DENV-2 (*K_d_* = 0.10 nM) demonstrate that discrete structural determinants are functionally conserved across these three viruses^19^. The region containing this epitope may be sufficient to allow the reactivation of memory B cells by heterologous NS1 proteins, despite the moderate level of global sequence identity between YFV and ZIKV/DENV NS1. This possibility, combined with the observation that antibodies recognizing this site bind with high affinity to the NS1 protein of several orthoflaviviruses, highlights the potential relevance of structurally conserved sites in shaping secondary immune responses. Further functional studies will be required to determine how these shared determinants contribute to cross-recognition and whether they influence protection during sequential *Orthoflavivirus* infections.

This structural mechanism contextualizes our observations in the ZIFAG cohort, where acute ZIKV infection triggered a marked increase in anti-YFV NS1 IgG that paralleled the rise in anti-ZIKV NS1 IgG levels. While classical linear epitopes common to the NS1 proteins of these two viruses are absent ^37^ - leading previous studies to consider these humoral responses as largely non-cross-reactive ^35,38^ - our findings suggest that conformity at the quaternary or structural level is sufficient to drive this cross-reaction. A similar pattern was observed for anti-DENV NS1 IgG, exclusively in individuals with prior DENV exposure, with DENV-naïve participants remaining negative.

Our data support the hypothesis that NS1-specific memory B cells elicited by prior YFV or DENV exposure can be recall during acute ZIKV infection through “original antigenic sin” mechanisms. These responses do not contribute to neutralization, but they may reflect immune memory imprinted by prior *Orthoflavivirus* exposure and highlight the immunological complexity in populations exposed to multiple *Orthoflavivirus* infections and/or vaccinations.

Our study has several limitations. First, the small number of patients with a history of DENV infection (*n* = 9) limits the robustness of our conclusions, as statistical analyses may be sensitive to outliers. Additionally, the sampling timepoint, 28 days after the onset of Zika symptoms was chosen to allow the detection of IgM, specific IgG, and virus-neutralizing antibodies. This may not correspond to the peak of the immune response for all detection methods ^27^ but YFV-specific memory B cells have been shown to be readily detectable by day 28 ^42^ and seroneutralizing antibodies have been shown to be detectable by day 14 ^20^. The DENV serotype responsible for prior infection could not be determined precisely because the available diagnostic tools lack specificity. We also had no information about the dates of prior DENV infections or YFV vaccinations, precluding any analysis of the potential influence of the time intervals since these prior events on the magnitude of NS1-specific IgG or neutralizing responses. Infections occurring shortly before ZIKV infection may have induced nonspecific antibody responses that might have skewed determinations. NS1-specific IgG antibodies are generally considered virus-specific but the ZIKV NS1 protein may contain epitopes common to the NS1 proteins of other orthoflaviviruses, including DENV ^43,44^. We cannot, therefore, rule out the possibility that some of the anti-DENV NS1 IgG detected in DENV-naïve individuals reflects cross-reactivity. The presence of this conserved epitope suggests a potential mechanism through which heterologous NS1 proteins could contribute to the recall of pre-existing memory B cells. However, the present study does not directly demonstrate that this epitope is involved in the observed antibody responses, and further functional studies will be required to address this question. Moreover, the absence of cellular samples from the ZIFAG cohort did not permit a strong mechanistic support of the memory recall hypothesis. Finally, another limitation is that ZIKV and YFV tend to be co-endemic in the regions in which they circulate. It is, therefore, difficult to identify individuals exposed to one virus who have not already been exposed to the other.

In summary, our results suggest that part of the humoral immune response directed against the NS1 protein of the ZIKV virus originates from an anamnestic response triggered by prior exposure to YFV or DENV. Such responses do not mediate virus neutralization but they highlight the specificity and imprinting of NS1-directed humoral memory shaped by prior *Orthoflavivirus* exposure. NS1 is involved in numerous pathological processes ^45^. Antibodies directed against this antigen are correlated with a lower incidence of pathological manifestations ^12,46^. A broader NS1-specific IgG immune response encompassing multiple orthoflaviviruses would have potential implications for disease progression, serological interpretation, and the feasibility of developing immune boosters against multiple orthoflaviviruses through natural infections and/or vaccinations with a live attenuated *Orthoflavivirus*.

## Data Availability

All data produced in the present study are available upon reasonable request to the authors

De-identified and aggregated data supporting the findings of this study are available from the corresponding author upon reasonable request. Individual-level data are not publicly available due to ethical and privacy restrictions.

## Acknowledgments

We thank the ISMEV platform (Immuno-Serological Monitoring of Emerging Viruses) for providing access to the equipment and technical resources used in this study. We thank all participants from the Vaxinnate cohort team, including Dr Alexandre Roulaud and the medical unit of the *21^ème^ Régiment d’Infanterie de Marine*, Fréjus, France.

## Authors’ contributions

S.M.: Conceptualization, Methodology, Formal Analysis, Investigation, Data Curation, Writing Original Draft,

N.A.: Investigation

K.M.: Investigation

E.L.D.: Methodology, Resources, Investigation, Data Curation

S.A.: Resources, Investigation

E.N.: Investigation

G.G.: Validation, Resources, Project Administration

A.T.: Investigation

B.C.: Conceptualization, Resources, Writing Original Draft, Funding Acquisition

M.M.: Conceptualization, Methodology, Resources, Funding Acquisition

S.B.: Conceptualization, Validation, Resources, Data Curation, Writing Original Draft, Funding Acquisition

C.B.: Conceptualization, Validation, Methodology, Formal Analysis, Data Curation, Writing Original Draft, Project Administration, Supervision, Funding Acquisition

All the co-authors were involved in reviewing and editing the draft

