## Supplementary material for "Kinetics and specificity of orthoflaviviruses NS1 IgG after Zika virus infection in Yellow fever and dengue immunized individuals: a memory recall hypothesis": Sup data

Supplementary data

|  | **DENV1 NS1** | **DENV2 NS1** | **DENV3 NS1** | **DENV4 NS1** | **ZIKV NS1** | **YFV NS1** |
| --- | --- | --- | --- | --- | --- | --- |
| **Number of patients** | 110 | 110 | 110 | 110 | 110 | 33 |
| **Minimum (MFI)** | 25 | 13 | 21 | 17 | 19 | 19 |
| **25% Percentile (MFI)** | 84 | 202 | 51 | 71 | 76 | 60 |
| **Median (MFI)** | 119 | 328 | 67 | 101 | 120 | 109 |
| **75% Percentile (MFI)** | 182 | 648 | 99 | 174 | 186 | 228 |
| **Maximum (MFI)** | 951 | 4059 | 479 | 1090 | 4939 | 1672 |
| **Range (MFI)** | 926 | 4046 | 458 | 1073 | 4920 | 1653 |
| **Mean (MFI)** | 163 | 556 | 86 | 157 | 207 | 239 |
| **Standard deviation (MFI)** | 88 | 415 | 40 | 99 | 158 | 222 |
| **Mean + 3 SD (MFI)** | 427 | 1802 | 207 | 453 | 679 | 903 |
| **Falses positives (%)** | 5 | 5 | 5 | 6 | 3 | 9 |

Supplementary table 1. **Intensity of the IgG response obtained with sera from negative patients**

|  | **Target** | **R^2^** | **a** | **b** | **c** | **Day max** | **Amp max (MFI)** |
| --- | --- | --- | --- | --- | --- | --- | --- |
| Without Pr DENV | DENV1 NS1 | 0,050036 | 315,81 | 0,25736 | 0,00089808 | 287 | 1047 |
|  | DENV2 NS1 | 0,069931 | 343,97 | 0,4 | 0,0028891 | 138 | 1657 |
|  | DENV3 NS1 | 0,029976 | 134,26 | 0,41496 | 0,0034538 | 120 | 647 |
|  | DENV4 NS1 | 0,05989 | 140,97 | 0,48804 | 0,0025669 | 190 | 1121 |
|  | ZIKV NS1 | 0,36522 | 2768,9 | 0,38948 | 0,0033076 | 118 | 12016 |
|  | YFV NS1 | 0,082861 | 2246,4 | 0,25894 | 0,0063428 | 41 | 4531 |
| with Pr DENV | DENV1 NS1 | 0,031147 | 894,95 | 0,3834 | 0,0051019 | 75 | 3195 |
|  | DENV2 NS1 | 0,041882 | 1087,1 | 0,42289 | 0,0052524 | 81 | 4556 |
|  | DENV3 NS1 | 0,028653 | 686,01 | 0,39323 | 0,0054879 | 72 | 2484 |
|  | DENV4 NS1 | 0,032246 | 1000,2 | 0,34842 | 0,003936 | 89 | 3366 |
|  | ZIKV NS1 | 0,30646 | 4132 | 0,31514 | 0,0031198 | 101 | 12911 |
|  | YFV NS1 | 0,014226 | 2503 | 0,10895 | 0,0022091 | 49 | 3432 |

Supplementary table 2. **IgG NS1 response, modeled by Wood’s equation, and parameters (a, b and c) used to determine the kinetics characteristics**

A

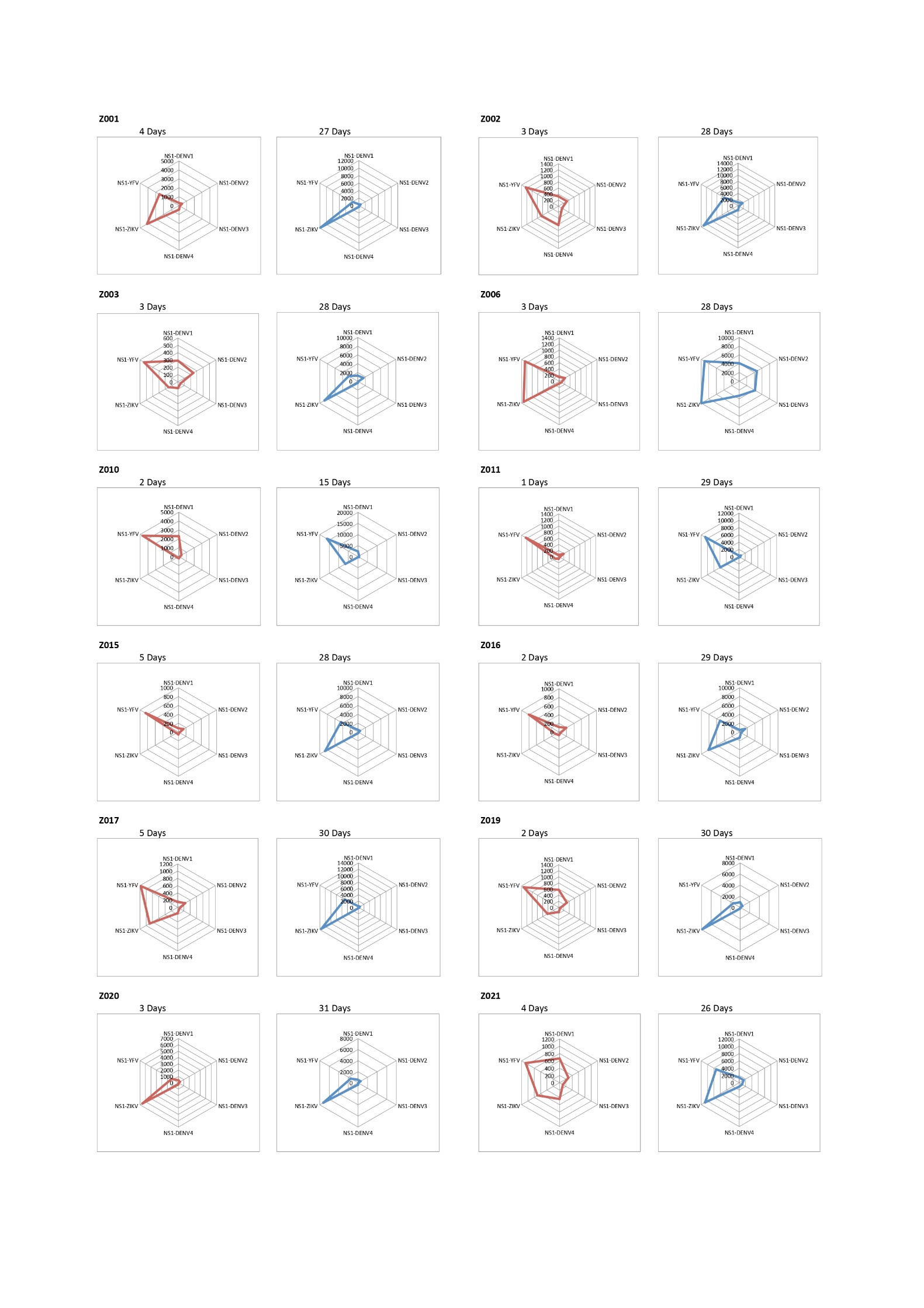

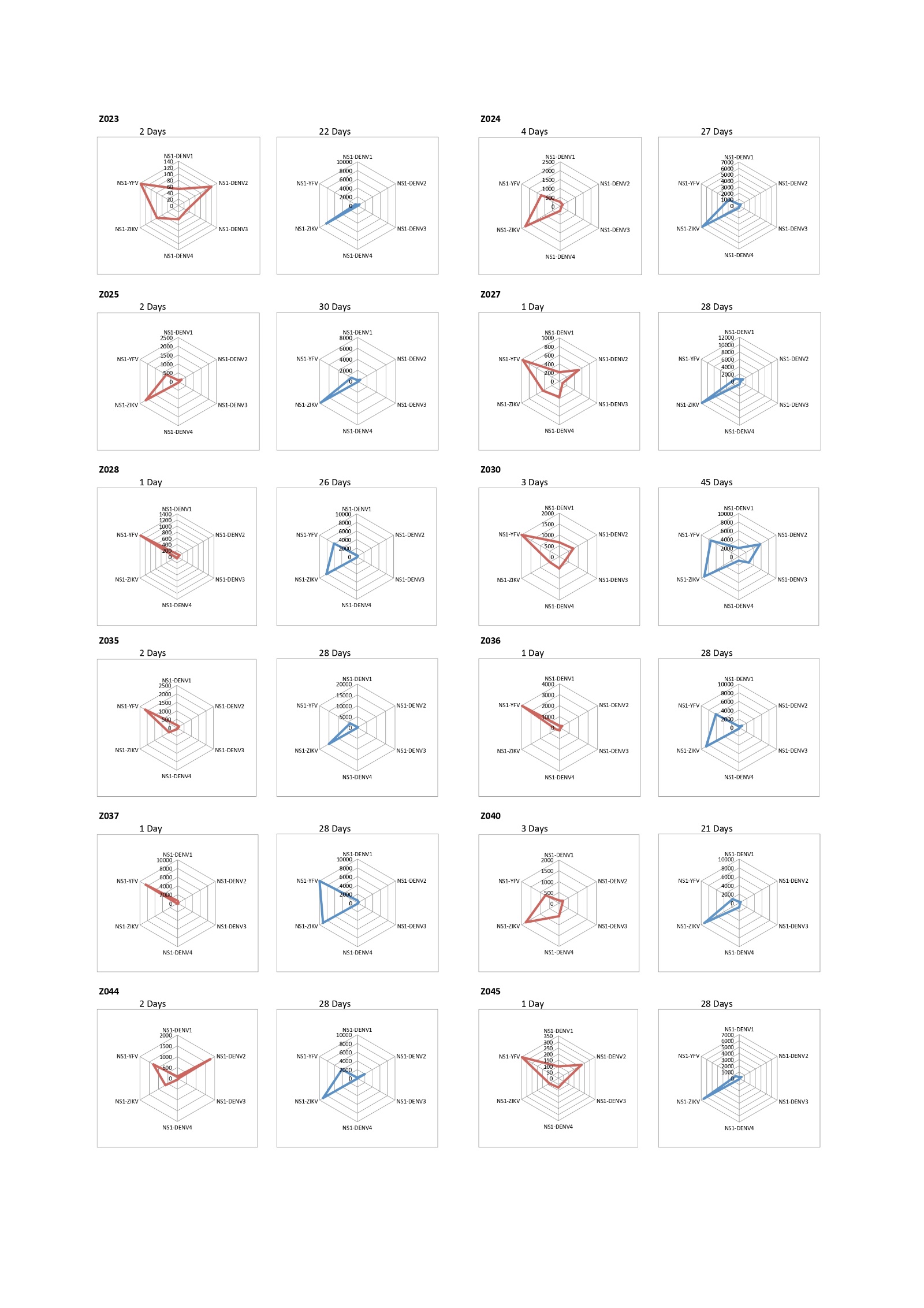

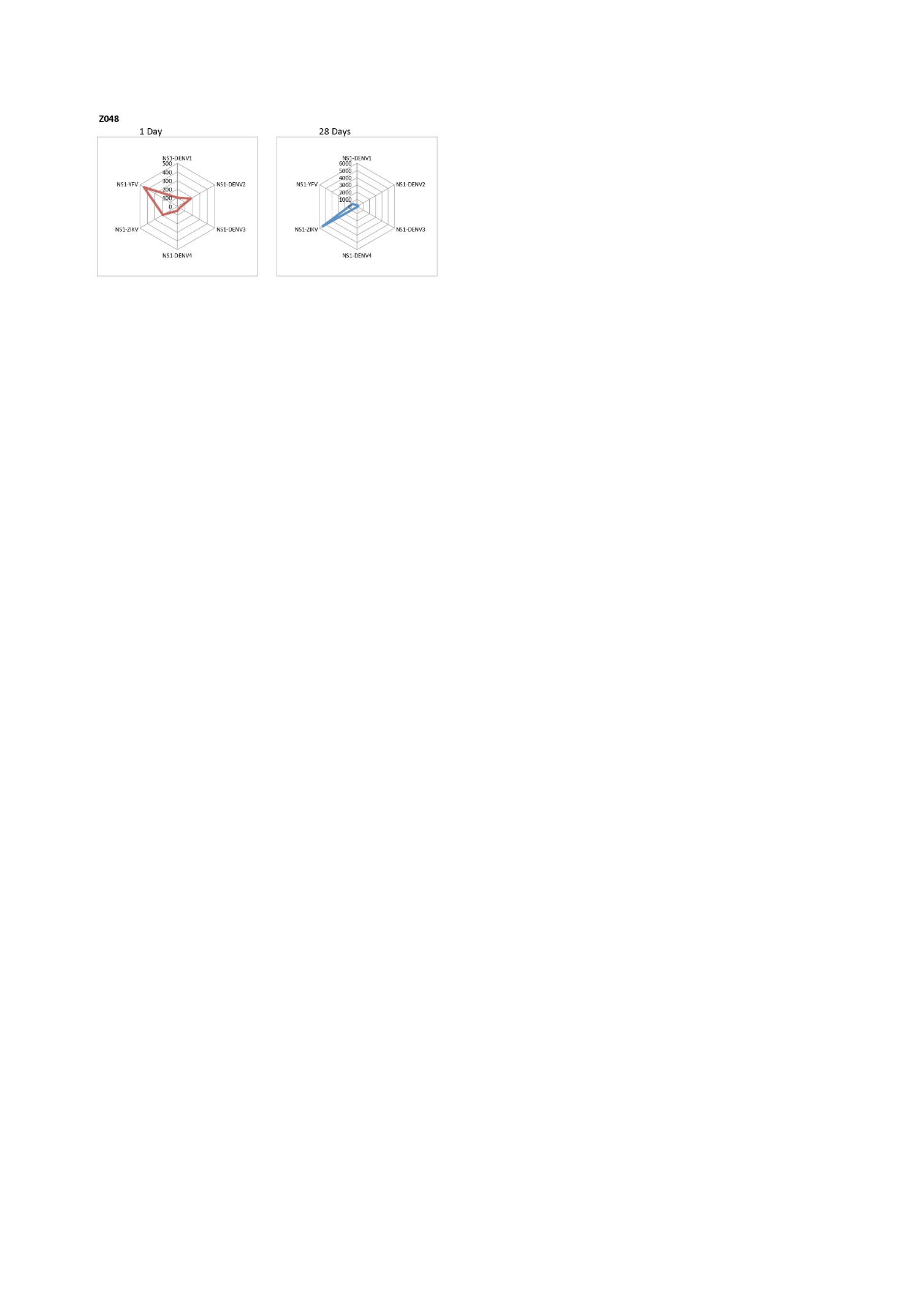

B

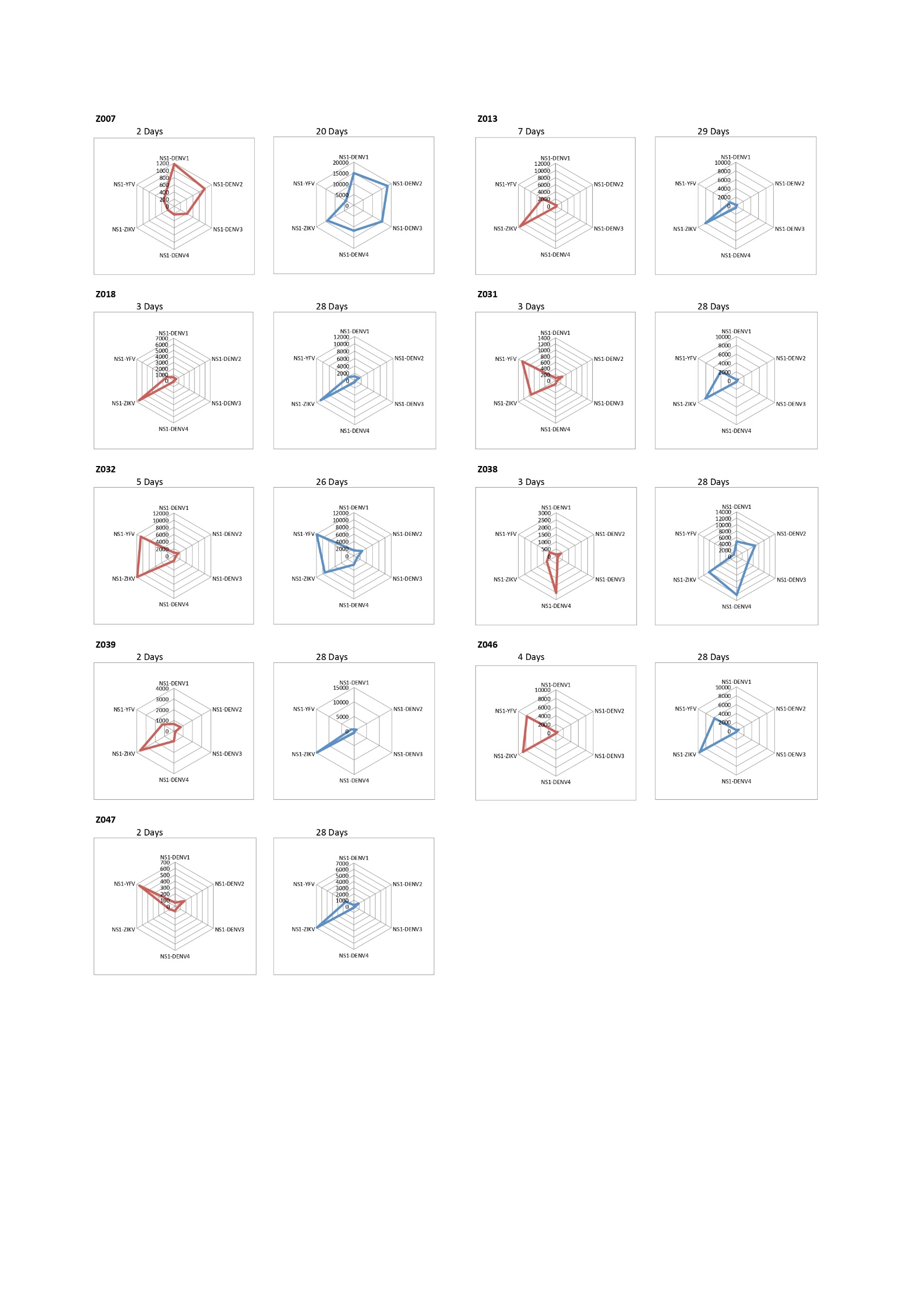

Supplementary Figure 1. **Evolution of IgG responses against *Orthoflavivirus* NS1 antigens in patient without prior DENV infection** (A) and with prior DENV infection (B), assessed at early and a later times after ZIKV infection.

The Radar plots display the median fluorescence intensity (MFI) of IgG responses targeting the NS1 proteins of ZIKV, YFV, and the four serotypes of DENV. Measurements were performed on the first sample collected after symptom onset and approximately one-month after symptom onset.

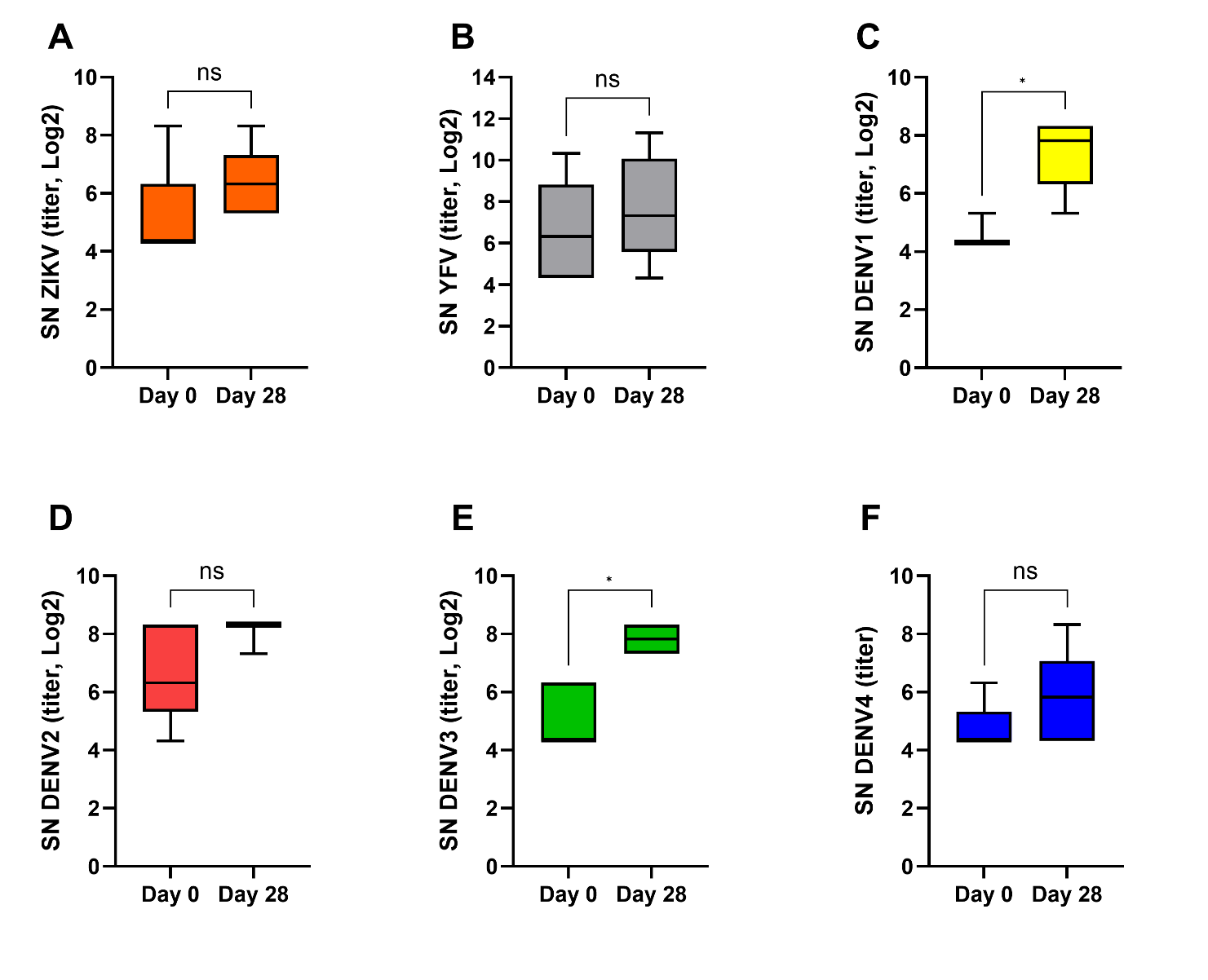

Supplementary Figure 2. **Seroneutralization (SN) measured on day 0 and 28 for Zika virus, yellow fever virus, and the four dengue virus serotypes in patients with a history of DENV infection**. Panels: A : ZIKV, B : YFV, C : DENV1, D : DENV2, E : DENV3, F : DENV4. The central line indicates the median and the whiskers represent the 10th and 90th percentiles.
